# Regional COVID-19 spread despite expected declines: how mitigation is hindered by spatio-temporal variation in local control measures

**DOI:** 10.1101/2020.07.17.20155762

**Authors:** Nicholas Kortessis, Margaret W. Simon, Michael Barfield, Gregory Glass, Burton H. Singer, Robert D. Holt

## Abstract

Successful public health regimes for COVID-19 push below unity long-term global *R*_*t*_ –the average number of secondary cases caused by an infectious individual. Most assessments use *local* information. Populations differ in *R*_*t*_, amongst themselves and over time. We use a SIR model for two populations to make the conceptual point that even if each locality averages *R*_*t*_ < 1, the overall epidemic can still grow, provided these populations have asynchronous variation in transmission, and are coupled by movement of infectious individuals. This emergent effect in pandemic dynamics instantiates “Parrondo’s Paradox,” -- an entity comprised of distinct but interacting units can behave qualitatively differently than each part on its own. For effective COVID-19 disease mitigation strategies, it is critical that infectious individuals moving among locations be identified and quarantined. This does not warrant indiscriminate prevention of movement, but rather rational, targeted testing and national coordination.

## Introduction

> “… *we’re a large country that has outbreaks in different regions, different states, different cities, that have different dynamics, and different phases*…”, (Dr. Anthony Fauci, quoted on CNN, 23 April 2020).

To control COVID-19, public health policy must drive the average effective net reproduction number (*R*_*t*_: the average number of secondary cases produced by transmission from a primary case at time *t*) below unity. Yet this statement applies *globally*. Local governments (e.g., states) craft policies based on *local* trends. In the USA (versus say New Zealand), policies are not coordinated across polities (e.g., states), and local control efforts wax and wane over time. New York (for instance) surges while Florida does not, but later this pattern flips, generating repeated waves of outbreaks varying among locations.

Tragically, spatial and temporal heterogeneity, coupled with movement of infectious individuals, can maintain global average *R*_*t*_ > 1, despite average *R*_*t*_ < 1 in all populations. This counterintuitive effect instantiates “Parrondo’s Paradox,” which describes how the behavior of complex systems with multiple interacting parts can differ qualitatively from the behavior of any single part (1). Parrondo’s Paradox occurs in population ecology (2-4), as well as physical, biological and financial systems (1, 5, 6) – but has not received attention in epidemiology. We illustrate the potential for this Paradox in the pandemic using a model (see Materials and Methods) which, though simple, captures essential elements of more complex, realistic models. We demonstrate that infectious individuals moving among locations with identical – but out of phase – temporal variation in transmission, sufficient for local control of COVID-19 when those locations are isolated, can drive global spread.

## Results

Figure 1 shows two typical scenarios where the disease cycles due to changing transmission rates, but nonetheless declines in each population when isolated (local 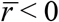; average *R*_*t*_ < 1). Without movement, the disease disappears. With movement and synchronous changes in transmission, the same holds (Fig. 1a,c). But with poor coordination across populations (viz., asynchrony), the disease spreads (Fig. 1b,d). Merely changing the relative timing of local control reverses global outcomes. Either synchrony *or* no movement between populations is sufficient for periodic enactment of social distancing policies to drive the disease extinct. However, with movement and asynchrony, these policies fail.

**Figure 1.**
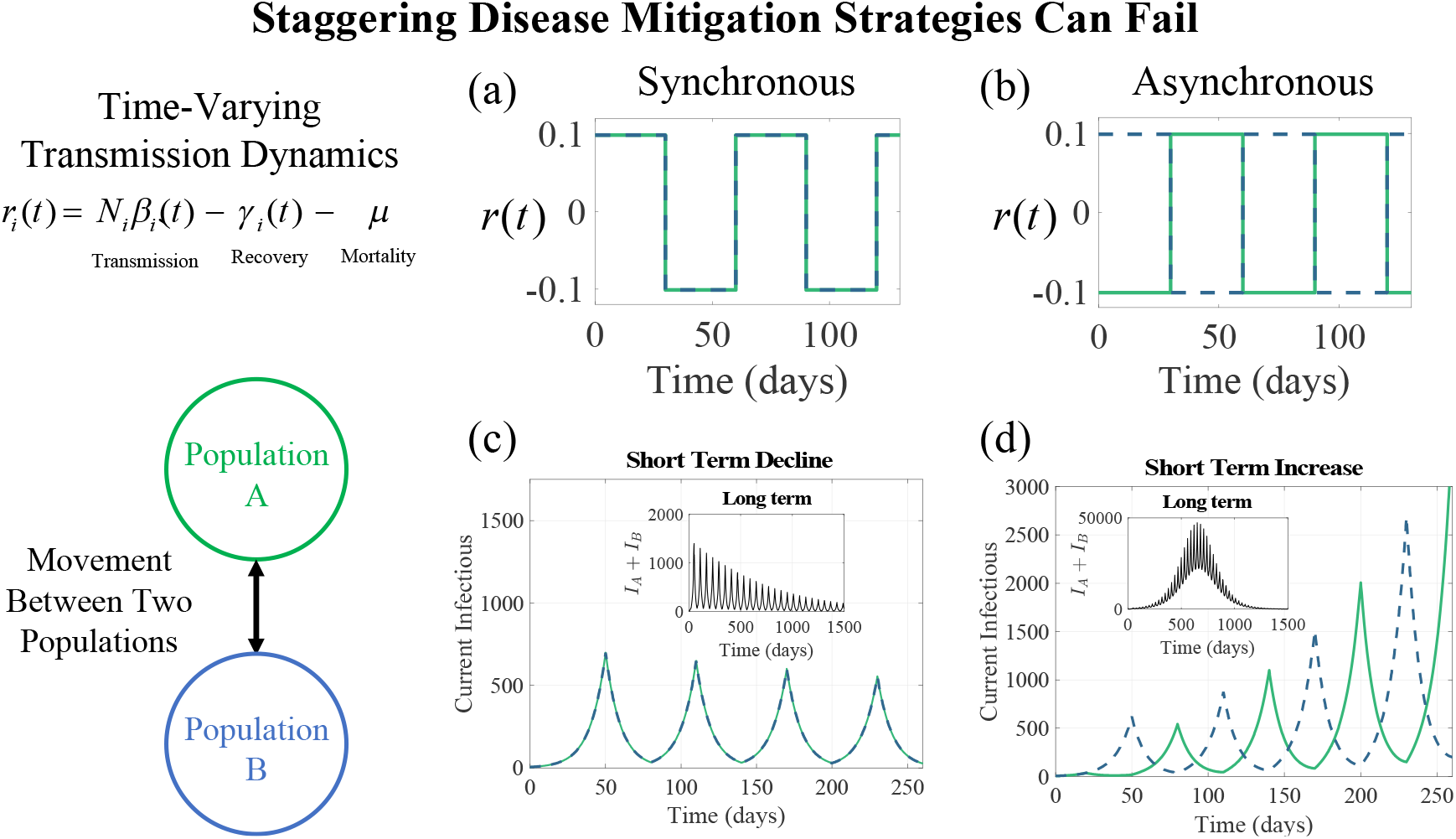
Movement and population differences in mitigation timing reverses disease eradication efforts. Transmission dynamics in two populations follow a square-wave function alternating between increasing prevalence (*r* > 0; *R*_*t*_ > 1) and declining prevalence (*r* < 0; *R*_*t*_ < 1) with 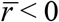 (average *R*_*t*_ over cycle < 1) (a) Pattern of transmission dynamics for (a) synchronous and (b) asynchronous populations. (c) Prevalence declines each cycle for synchronous populations, but (d) increases for asynchronous populations. Insets show long-term dynamics. Populations are identical except for timing of parameter shifts.

Movement and asynchrony cause long-term harm when combined, as illustrated in Figure 2. Model parameters are conservative with respect to estimates of SARS-CoV-2 dynamics (7-9) (see Materials and Methods). Deaths, total cases, and peak case numbers increase with movement and asymmetry (Fig. 2a-c), with potentially devastating consequences. For example, increasing movement from 0.1% to 0.5% for mostly asynchronous populations increases cases and deaths nearly an order of magnitude (light green curve, Ω = 0.75; Fig. 2c).

**Figure 2.**
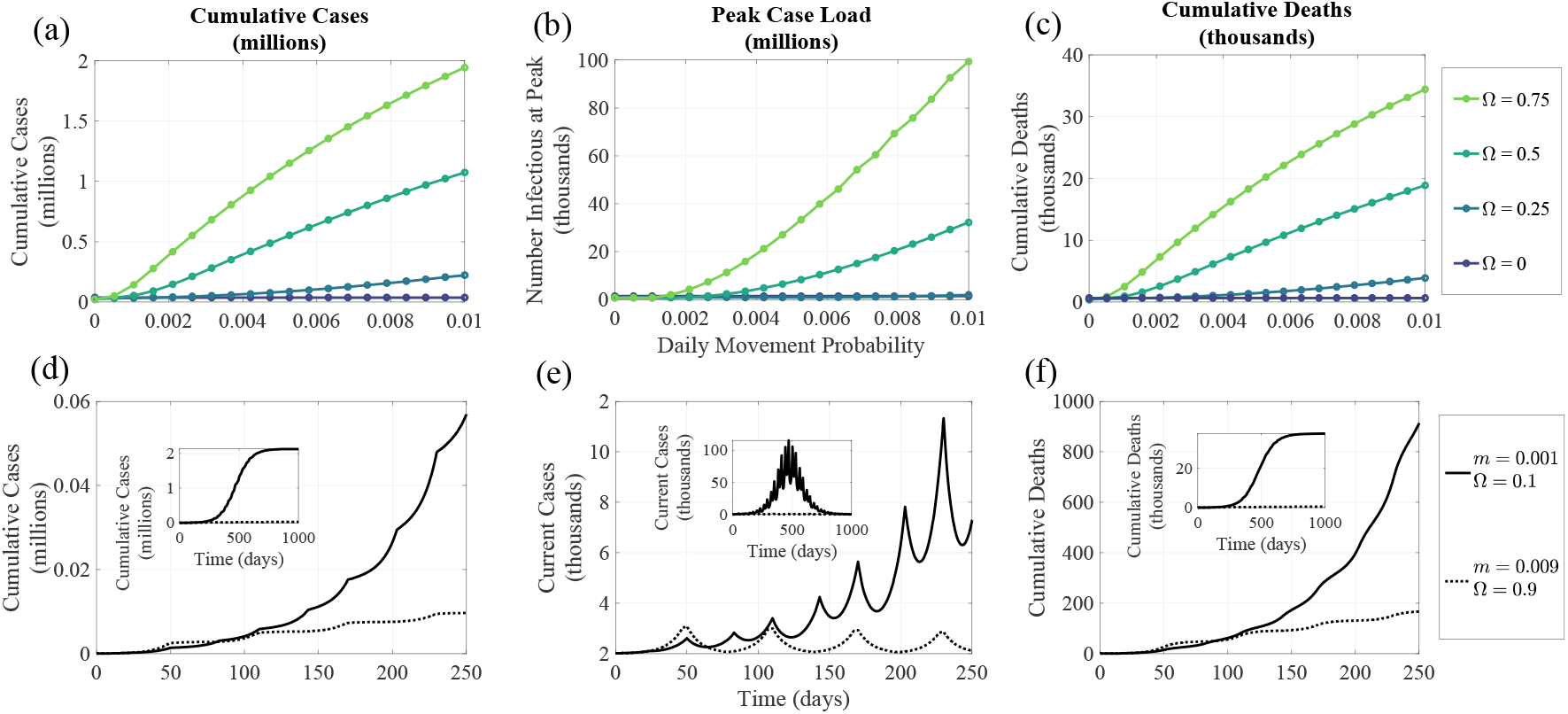
Consequences of Parrondo’s Paradox for disease outcomes. (a) Cumulative infections at the end of the epidemic, (b) peak infectious, and (c) total deaths at the end of the epidemic. Dynamics of (d) cumulative cases, (e) current infectious, and (f) cumulative deaths on short (main) and long (insets) timescales (dotted line not visible in insets due to comparatively small values).

The synergistic effects of movement and asynchrony emerge quickly in an epidemic. Disparities between a scenario with low movement and asynchrony (dotted lines, Fig. 2d-f) and one with moderate movement and asynchrony (solid lines, Fig. 2d-f) accumulate early on, with substantial human, economic, and public health impacts.

## Discussion

We suggest a version of Parrondo’s Paradox could arise in the current pandemic. While spatial processes are increasingly recognized as critical in epidemiological theory (e.g., the evolution of resistance by nosocomial pathogens, (10)), as is temporal variation in disease transmission (e.g., fluctuations in *R*_*t*_ driven by social factors such as conflict (11)), their combined effect is underappreciated. Together, spatial and temporal processes can have powerful and surprising effects on disease dynamics, with devastating consequences. This result requires only two conditions: 1) disease management strategies in different populations do not coincide in time, and 2) some infectious individuals move between populations. Restricting movement among populations and increasing coordination can reduce disease impacts.

Our goal is to illustrate a qualitative effect, not predict specific disease outcomes; many real-world complexities need consideration for concrete application of our insights to the COVID-19 pandemic (e.g., (7-9)). However, we expect the qualitative effect pertains to realistic scenarios, as known for Parrondo effects in spatial population ecology (4).

Our findings suggest integrated policy actions at regional, national, and global scales:

1. Intensify mitigation locally (reduce 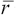 and prolong social distancing).
2. Coordinate strategies (synchronize the timing of lockdown and re-opening).
3. Reduce movement of infectious individuals.

The impact of these actions is contextual, because their dynamical effects are intertwined. Assessing the potential for Parrondo’s Paradox to affect the COVID-19 pandemic using realistic, spatially structured, and necessarily complex epidemiological models (12), we believe, should be carried out, and soon. A holistic, multi-regional and dynamical perspective is absolutely essential for effective control of this pandemic.

## Materials and Methods

### SIR Dynamics

Two populations (*i* = *A,B*) each follow a time-varying SIR model:

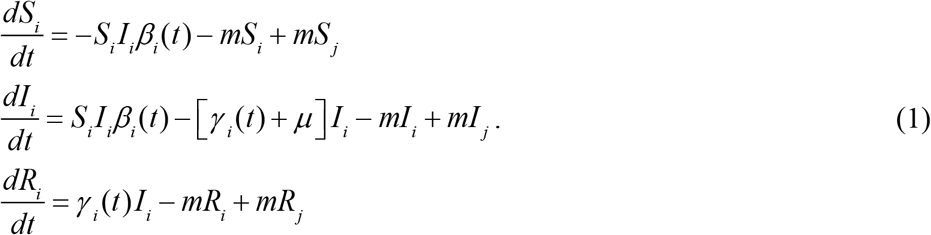

*S*_*i*_, *I*_*i*_, and *R*_*i*_ are local susceptible, infectious, and recovered individuals, respectively (local population size is *N*_*i*_, assumed 5 million for *i* = *A,B*). *β*_*i*_(*t*) = transmission rate, *γ*_*i*_(*t*) = recovery rate, *μ* = disease mortality rate, and *m* = per-capita movement rate. When COVID-19 is rare, *S*_*i*_ ≈ *N*_*i*_, and disease spread follows

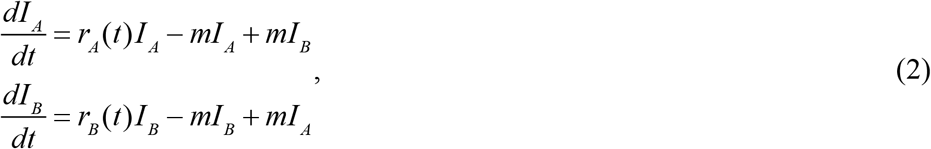

where *r*_*i*_(*t*) = *N*_*i*_*β*_*i*_(*t*) – *γ*_*i*_(*t*) – *μ* is the local instantaneous per-capita rate of change in the infectious class without movement [note *R*_*i*_(*t*) = *N*_*i*_*β*_*i*_(*t*)/(*γ*_*i*_(*t*) + *μ*)]. *I*_*i*_ increases for *r*_*i*_(*t*) > 0 (*R*_*t*_ > 1) and declines for *r*_*i*_(*t*) < 0 (*R*_*t*_ < 1). We assume square-wave alternation in *r*_*i*_(*t*) between high and low values, with period *T* (assumed 60 days; Fig. 1). Growth is positive (*rN* > 0) for half the period (*N* denotes “normal,” with typical contact patterns) and negative (*rS* < 0) otherwise (*S* indicates “social distancing,” with reduced contact), resembling shifts in policy and behavior. Change in *I* over each cycle follows 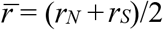; importantly, we assume temporal mean 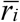 is negative. Populations only differ in timing of switches between states, with asynchrony Ω defined as the fraction of time *rA*(*t*) ≠ *rB*(*t*) (0 ≤ Ω ≤ 1).

Parameter values for normal (and empirical estimates for reference) are (for normal and social distancing states, respectively): *βN* = 0.1988 and 0.0288 day^-1^ (0.35-1.12, ref (8)), *γ* = 0.098 and 0.128 day^-1^ (0.29, ref (8); 0.07, ref (9)), and *μ* = 0.002 (0.04; ref (9)). Thus, *rN* = 0.0988 (0.17-0.23, ref (7)), *rS* = −0.1012, and 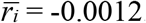. Case doubling time is ln(2)/*rN* = 7.02 days (3-4 days, ref (7)) and case halving time is ln(1/2)/*rS* = 6.85 days. *R*_*t*_ = 1.988 and 0.2215 for *rN* and *rS*, respectively (2.4-4, refs (7-9)).

### Simulating dynamics

We numerically solved the model (eq.1) in Matlab 2019b. All simulations began with 5 infections in each population. The infectious class grew exponentially for 20 days at *rA* = *rB* = *rN* to simulate disease emergence and then square-wave cycling began. To evaluate long-term dynamics, we simulated dynamics for 100 cycles.

### Measuring Epidemiological Outcomes

Cumulative case numbers, cumulative deaths, and prevalence were calculated from numerical solutions to the model. We estimated peak prevalence (maximum value of *I*_*A*_(*t*) + *I*_*B*_(*t*)) by solving equation (1) over 2,000 evenly spaced time units per cycle (i.e., every 0.03 days). Cumulative cases each time step are ∑_*i*={*A,B*}_*N*_*i*_ – *S*_*i*_(*t*), and cumulative deaths are ∑_*i*={*A,B*}_*N*_*i*_–[*S*_*i*_(*t*)+*I*_*i*_(*t*)+*R*_*i*_(*t*)]. Movement follows a Poisson-point process with rate *m*, implying the probability of moving in one timestep is 1 – exp(–*m*).

## Data Availability

The manuscript is a modeling story, all parameters used in the model are drawn from papers cited in the manuscript.

## Acknowledgments

This work was funded by USDA (award number 2017-67013-26870) as part of the joint USDA-NSF-NIH Ecology and Evolution of Infectious Diseases program, the National Science Foundation (DEB-1655555), and the University of Florida Foundation. Derek Cummings provided helpful advice as did Alison Galvani and Ira Longini.

